# Development, validation and clinical implementation of cellular swab materials for quality assurance of pharmacogenomic point-of-care testing

**DOI:** 10.64898/2026.10.13.26365321

**Authors:** Henry M Thomas, John H McDermott, William G Newman, Shaun Ainsworth, Gino Miele, Simon J Patton, Rosa Morra

**Affiliations:** EMQN CIC, 3rd Floor, ICE Building, Exchange Quay, Salford, M5 3ED, UK; Manchester University NHS Foundation Trust, Manchester Centre for Genomic Medicine, Manchester University NHS Foundation Trust, Manchester, UK; Division of Evolution, Infection and Genomics, School of Biological Sciences, Faculty of Biology, Medicine and Health Sciences, University of Manchester, Manchester, UK; Genedrive PLC, 48 Grafton St, Manchester, M13 9XX, UK

## Abstract

**Background:** Pharmacogenomic point-of-care testing (PGx POCT) enables genotype-informed prescribing at the bedside, but implementation is limited by the lack of suitable reference materials for clinical verification and standardised external quality assessment (EQA).

**Methods:** Fixed lymphoblastoid cell lines were formulated as swab-based materials at defined cellular inputs for *MT-RNR1* POCT. Materials were evaluated using Genedrive® assays, quantitative PCR, acceptance quality limit batch release, workflow-perturbation testing and stability studies, then deployed for clinical verification and in an international EQA programme.

**Results:** Materials were homogeneous, workflow compatible and reproducible, with 100% correct genotype calls at the lowest intended manufacturing cell load. Clinical verification of 11 Genedrive® systems across five neonatal units demonstrated 99% concordance with expected genotype classifications (232/234 results). Seven EQA surveys yielded 253 participant results, with full-mark rates of 87.5–100%, and six test failures. Most failures occurred with low cell-input materials near the assay limit of detection. The materials supported genotype concordance, performance monitoring and identification of workflow and sample-related variability.

**Conclusions:** Workflow-compatible cell-based reference materials combined with EQA provide a scalable framework for quality-assured PGx POCT beyond specialist laboratories. Clinical verification of Genedrive® systems within neonatal units using the cell-based swab materials, together with deployment of an International MT-RNR1 PGx POCT EQA programme, demonstrated their suitability for supporting both pre-implementation testing and ongoing quality assurance in decentralised healthcare settings. The framework may provide a model for the implementation, clinical verification and ongoing quality assessment of PGx POCT services. Further evaluation across platforms, analytes, settings and participant groups is required to establish broader applicability.

## Introduction

Pharmacogenomics (PGx) is a central component of precision medicine, enabling the tailoring of drug therapy according to an individual’s genetic profile, with the potential to improve treatment efficacy and reduce adverse drug reactions. Despite significant technological advances and the increasing availability of clinically actionable genetic information, the integration of pharmacogenomic testing into routine clinical practice remains inconsistent and limited by a range of implementation barriers (1,2). These include challenges related to clinical workflows, education, interpretation of results, clinically relevant turnaround time and the lack of standardised approaches to testing and reporting (3,4). Turnaround time is particularly important when prescribing decisions must be made rapidly, making conventional laboratory-based testing impractical for some clinical pathways (5). PGx Point-of-Care Testing (POCT) has emerged as a promising strategy to address the delays associated with centralised laboratory workflows and prolonged turnaround times. A key example is testing for the *MT-RNR1 1555A>G* variant, which predisposes to aminoglycoside-induced hearing loss (6). Aminoglycosides are commonly used in the newborn setting to treat severe infection, where results are required within an hour, a timeframe incompatible with conventional laboratory testing. As such, a *MT-RNR1* POCT testing strategy has been developed and evaluated for use in acute neonatal care (7). *CYP2C19* genotyping represents another clinically relevant use case, where rapid identification of loss-of-function alleles can inform antiplatelet therapy selection in acute cardiovascular pathways (8). The development and validation of rapid *CYP2C19* POCT assays capable of delivering actionable results within clinically relevant timeframes further highlight the potential of this approach to support genotype-guided prescribing when timely treatment decisions are required (9).

However, the safe clinical implementation of genomic POCT requires robust quality assurance systems, including external quality assessment, together with appropriate reference materials to support validation, verification and ongoing performance monitoring (10,11). Decentralised testing performed by non-specialist healthcare personnel, can introduce variability in sample handling, analytical performance, and result interpretation. In addition, decentralised testing settings often lack the harmonised quality management frameworks routinely available in accredited laboratories, making consistent performance across sites more difficult to assure. These challenges are particularly important in genomic testing, where errors may directly influence therapeutic decisions and patient outcomes.

EQA is a cornerstone of laboratory quality systems, providing independent evaluation of testing performance and helping to identify analytical and reporting errors (12). For POCT, EQA has additional considerations because participants may include non-laboratory healthcare professionals and EQA schemes should be designed to assess, where possible, the pre-examination, examination and post-examination phases of testing (13). Effective EQA relies on commutable reference materials that behave similarly to patient samples across different analytical platforms and workflows (14). In genomic POCT, this requirement extends beyond analytical performance to include the unique workflow characteristics of near-patient testing.

Conventional molecular quality control materials based on purified genomic DNA (gDNA) or synthetic oligonucleotides are poorly suited to genomic POCT, where cell lysis, nucleic acid extraction, amplification, and detection are typically integrated within a single workflow. Consequently, reference materials must mimic both the cellular composition and handling characteristics of clinical specimens. Patient-derived clinical specimens would be preferable in principle, but obtaining sufficient volumes of well-characterised positive material is often impractical, particularly for rare but clinically important variants such as *MT-RNR1* m.1555A>G (15). This limitation represents a major barrier to the safe and scalable implementation of genomic testing in decentralised settings. Here, we address this unmet need by developing contrived cell-based materials for *MT-RNR1* POCT applications. The materials were designed to mimic clinical buccal swab specimens and to provide workflow-relevant assessment of sample processing and genotype detection.and evaluating their use within an integrated EQA framework. We evaluated their analytical performance, reproducibility, homogeneity, stability, workflow robustness and suitability for clinical verification and EQA applications. We further demonstrate their deployment within an international *MT-RNR1* PGx POCT EQA programme, providing a practical framework for quality-assured implementation of pharmacogenomic POCT beyond specialist laboratory settings.

## Material & Methods

### Cell Line Generation

Cell lines were established to generate cell-based buccal swab materials containing the mitochondrial *MT-RNR1* m.1555A>G variant, a clinically significant pharmacogenetic biomarker associated with aminoglycoside-induced ototoxicity and the primary target of the Genedrive MT-RNR1 point-of-care test. Whole blood samples collected from individuals with previously confirmed pharmacogenomic results were processed at the Genomics Diagnostic Laboratory, St Mary’s Hospital (Manchester). Patients were consented and samples generated as part of the Manchester Gene Identification Consortium (MAGIC) study (Research Ethics Committee Reference 11/H1003/3). Samples underwent Epstein–Barr virus (EBV)-mediated transformation of B lymphocytes (16–18). Transformed cells were cultured in RPMI 1640 containing 2mM L-Glutamine and Penicillin/Streptomycin at 500 units each per ml, and either 20% or 10% Fetal Bovine Serum (R20 or R10 media) under standard conditions (37°C, 5% CO₂). Cell suspensions were expanded and aliquoted into sterile cryovials, each containing approximately 2 - 3 × 10⁶ viable cells per ml. Cell lines were cryopreserved in RPMI 1640 containing 2mM L-Glutamine and Penicillin/Streptomycin at 500 units each per ml, and 20% Fetal Bovine Serum (R20) plus 10% Dimethyl Sulfoxide (DMSO) and stored at −80°C prior for long-term storage.

### Cell line genotype confirmation

Sanger sequencing was performed to confirm the presence of target variants within the selected cell lines (Supplemental Method S1).

### Cell Line expansion and Fixation

Following expansion, cell cultures were harvested at optimal density to ensure high viability and consistent cellular composition. Cells were subjected to a fixation process to stabilise cellular structure and preserve genetic material for downstream applications. Fixation was performed using ThinPrep® PreservCyt® Solution (Hologic cat. 70406-002). This process renders cells non-viable while preserving nucleic acid integrity and structural characteristics, enabling their use as stable reference materials for molecular testing applications (19).

### Manufacturing of cell-based swab materials

Fixed cell suspensions were used to manufacture flocked swab-based materials designed to simulate clinical sample collection workflows. Target cell concentrations were informed by both the analytical sensitivity of the Genedrive® *MT*-*RNR1* assays and cell count measured in clinically derived buccal swab specimens. Assay-specific limits of detection (LoD) were converted to equivalent cell numbers per swab using the Genedrive® sample preparation workflow; however, as these values assume complete cell recovery and transfer, manufacturing targets were selected to span clinically relevant cellular inputs observed in patient samples while remaining above the minimum analytical detection threshold (Table 1). Swab manufacture was performed by Microbix Biosystems Inc. (Toronto, Canada) using validated proprietary processes to deposit fixed cells onto flocked swab matrix at predefined target cell loads. Manufacturing batches comprised a single production lot of at least 135 swabs of the same genotype and cellular input level. Swabs were manufactured to a target cell load within ±10% of the specified value. Manufactured swabs were individually packed in foil pouches and were stored at +4–18°C prior to distribution.

**Table 1.**
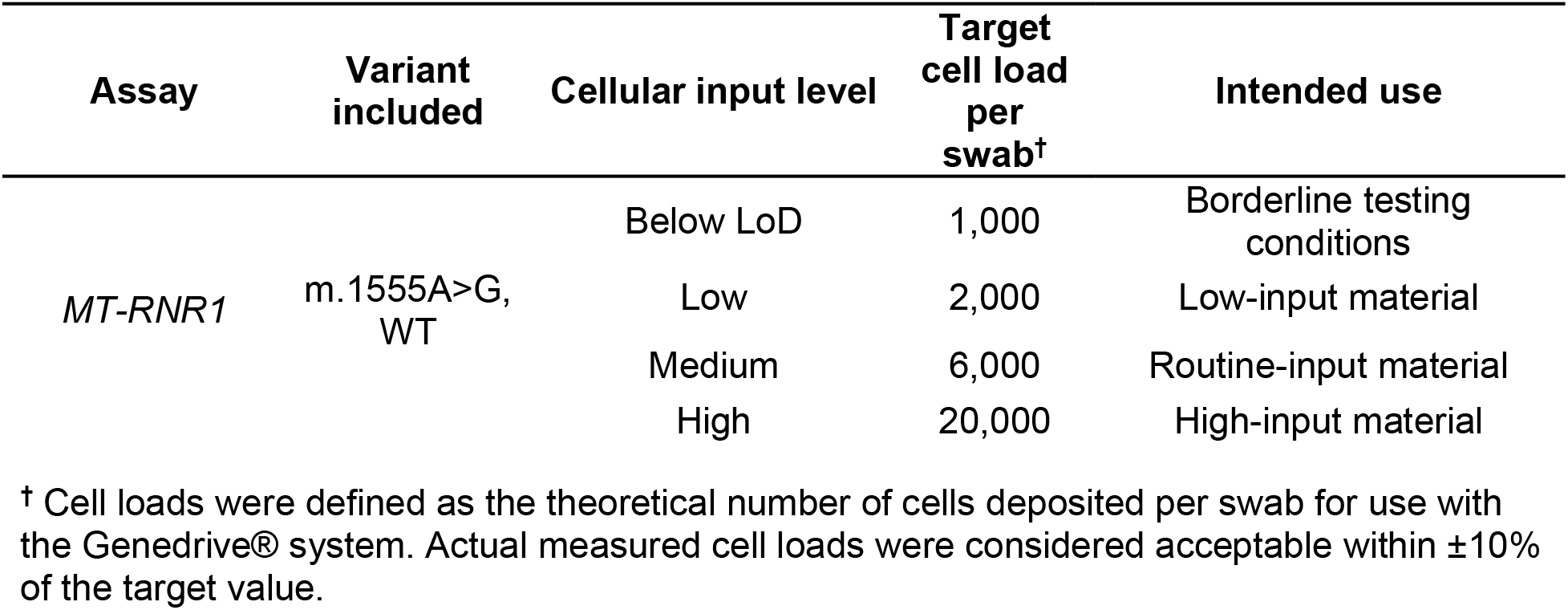
Pharmacogenomic swab materials manufactured for assay evaluation.

### In-house validation of reference materials

The performance of the cell-based swab materials was evaluated through in-house validation studies. Variant detection was assessed qualitatively using the Genedrive® *MT-RNR1* kit. These assays determine genotype through analysis of genotype-specific melting curve profiles generated following loop-mediated isothermal amplification (LAMP) of the *MT-RNR1* target region (20). Individual target peak height (RFU) was used as an indicator of assay signal intensity and cellular recovery, whilst melting temperature (Tm) positions were used to confirm genotype assignment. To confirm the cellular input delivered by the swabs, the amount of genomic DNA recovered from the materials was estimated by qPCR using a standard curve generated from serial dilutions a Human Genomic DNA (Sigma-Aldrich, Catalogue number 69237-3) (Supplemental Method S2). To further verify the intended difference in cellular input between the high and low-cell-load materials, a 10-fold dilution of the high-cell-load material was analysed using the Genedrive® assays and compared with the manufactured low-cell-load materials. This approach enabled estimation of DNA yield, inference of cellular input, and assessment of batch-to-batch consistency.

Validation studies assessed detection of the expected variants at or near the assay limit of detection, reproducibility across independent replicates, performance across different cell loads, and robustness to deliberate deviations from the assay instructions for use (IFU). Batch release testing was performed using AQL-based attribute sampling according to ISO 2859-1 (21) (equivalent to ANSI/ASQ Z1.4). A defect was defined as failure to generate a valid result or an incorrect genotype assignment. Testing was performed on the lowest cellular input material within each production batch, representing the most challenging manufacturing condition and the material most likely to reveal variability in cellular loading or assay performance. Initial *MT-RNR1* materials were evaluated using AQL Level II sampling plans. Following demonstration of manufacturing consistency, subsequent *CYP2C19* materials were assessed using AQL Level I sampling plans as a risk-based approach to reduce sampling requirements while maintaining confidence in product quality.

### Stability studies

Stability studies were performed to assess the integrity and performance of the manufactured swab materials during storage, handling, and transportation (Supplemental Method S3).

### Clinical Device verification package

The artificial swab materials were incorporated into a clinical device verification package comprising *MT-RNR1* wild-type and m.1555A>G specimens manufactured at both high and low cellular input levels. Accompanied by a standardised verification protocol (Supplemental Table S1), the package was designed to support site-level verification of Genedrive® *MT-RNR1* Systems in accordance with ISO 15189:2022 requirements (22) and CLSI EP12 principles for qualitative test evaluation (23). Primary verification of newly introduced systems used repeat testing by independent operators over multiple days, whereas secondary verification of additional systems used a reduced verification protocol (Supplemental Table S1). Verification outcomes were assessed based on concordance with expected genotype classification and occurrence of test failures.

### External Quality Assessment (EQA) scheme design

A pilot EQA scheme was established to evaluate the suitability of the developed *MT-RNR1* cell-based materials in a real-world setting. The EQA was conducted as part of the Pharmacogenetics to Avoid Loss of Hearing (PALOH-UK) implementation study (ISRCTN10216938), which focused on *MT-RNR1* point-of-care testing to guide aminoglycoside antibiotic prescribing. Participating laboratories received swab-based samples and analysed them using their routine *MT-RNR1* POCT workflows. During the first year of the pilot, six EQA surveys were distributed at approximately two-month intervals, with each survey comprising three samples(24). The EQA panel included samples with defined genotypes and varying cell loads, and participant results were assessed against predefined performance criteria.

## Results

### Design of workflow-compatible cell-based swab materials

A genotype-confirmed panel of lymphoblastoid cell lines (LCLs) was established for development of pharmacogenomic POCT reference materials. Cell lines carrying the *MT-RNR1* wild type and *m.1555A>G* variant, were selected to generate workflow-compatible cell-based swab materials for Genedrive® testing workflows (25,26). These characterised cell lines subsequently formed the basis of a scalable manufacturing process for cell-based swab materials suitable for clinical device verification and external quality assessment (EQA) applications.

### Manufacturing strategy and cell-load design

Cellular input levels comprised a cellular level which was below the assay limit of detection (LoD), together with low, medium and high-input materials spanning the range of specimen cellularity likely to be encountered in clinical practice (Table 1). This design enabled assessment of assay performance across both expected and borderline testing conditions while supporting evaluation of workflow robustness. The resulting material panel included both positive and negative genotype controls (Table 1) and was subsequently used to assess analytical performance, manufacturing consistency, stability and suitability for EQA applications.

### Analytical performance across intended and challenge cell loads

Initial analytical feasibility was evaluated prior to formal validation using a small set of manufactured *MT-RNR1* swab materials produced at the low cell-load manufacturing specification (Supplemental Table S2). All target variants were correctly identified using the corresponding Genedrive® assays, with 100% concordance observed between expected and reported genotypes across *MT-RNR1* materials (14/14 correct genotype calls; Supplemental Table S2). These findings confirmed that the selected low cell-load manufacturing specification provided sufficient cellular input for reliable genotype assignment across all target variants. As low cellular inputs close to the assay LoD present a greater analytical challenge, successful performance at this specification provided confidence that materials manufactured at higher cell-load specifications would also support robust genotype assignment.

Following this preliminary feasibility assessment, analytical sensitivity was evaluated across a broader range of cellular input levels, including conditions below the assay LoD, to characterise assay performance under increasingly challenging testing conditions. Swab materials manufactured at varying cellular input levels were tested using the corresponding Genedrive® assay. All samples containing high, medium and low cell loads produced the expected genotype result. In contrast, materials manufactured below the assay LoD exhibited an increased frequency of test failures and a greater proportion of assay target peak heights below the acceptance threshold, consistent with insufficient cellular input (Figure 1 and Supplemental Fig. S1). However, when sufficient amplification signal was generated, samples remained within the expected genotype-specific melting temperature (Tm) cluster and were assigned the correct genotype (37/40 samples; 92.5% across both genotypes), demonstrating that reduced performance was attributable to insufficient cellular input rather than impaired genotype discrimination (Supplemental Fig. S1).

**Figure 1.**
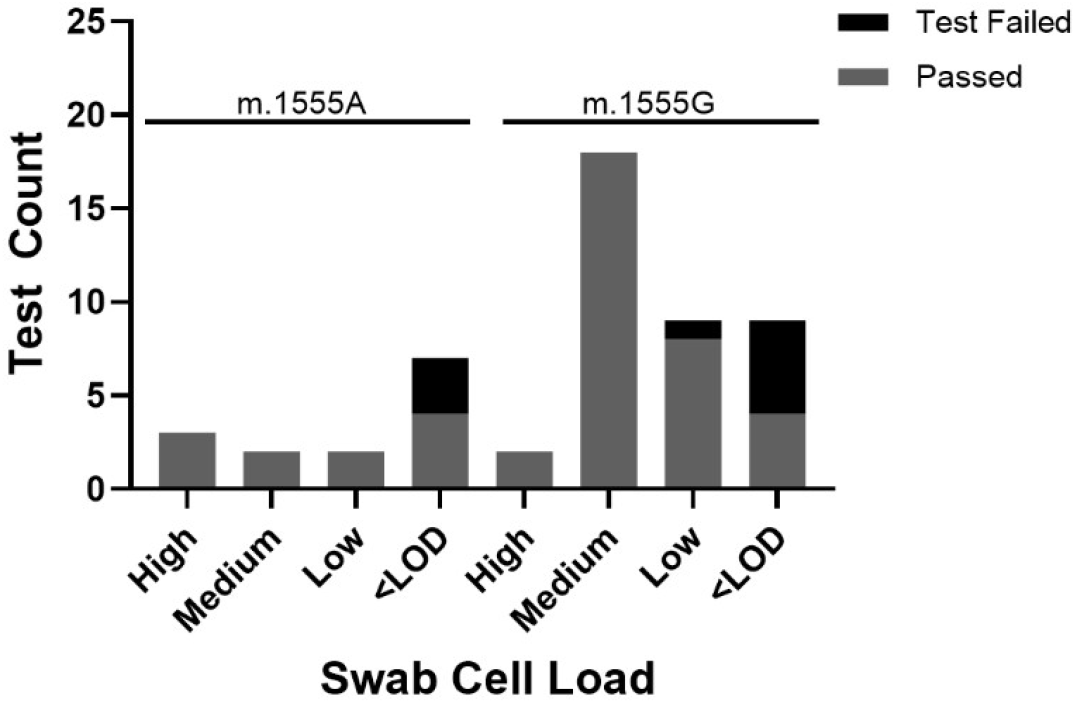
Analytical sensitivity study across a range of cell loads, including below LoD materials. Artificial swab materials containing decreasing cell loads were tested using the Genedrive® *MT-RNR1* ID Kit. Materials manufactured above the limit of detection produced the expected genotype calls 35/36 (97.2%), whereas materials manufactured below the assay limit of detection showed reduced performance producing the expected genotype 8/16 (50%). This reduction in performance was statistically significant (two-sided Fisher’s exact test, p = 0.0001).

Semi-quantitative qPCR using a genomic DNA standard curve further confirmed separation between high- and low-cell-load materials. For *MT-RNR1* materials, mean DNA recovery was 16.3 ± 4.7 ng/swab for high-cell-load materials and 1.8 ± 0.3 ng/swab for low-cell-load materials, corresponding to an approximately 9.1 ± 2.9-fold difference in recovered DNA (Supplemental Table S3). This closely matched the intended 10-fold difference in cellular input and provided independent verification of the manufacturing strategy used to generate distinct material levels. Functional confirmation of the intended cell-load difference was obtained by testing a 10-fold dilution of the high-cell-load material released in the assay buffer on the Genedrive® platform. The diluted material produced concordant genotype assignments and assay performance comparable to the corresponding manufactured low-cell-load material, demonstrating that the observed differences in assay performance were attributable to the intended variation in cellular input rather than manufacturing variability (Supplemental Table S3).

### Homogeneity, Reproducibility and Fitness-for-Purpose Assessment

Manufacturing consistency was assessed using complementary quantitative and qualitative approaches. Semi-quantitative qPCR was used to estimate recovered cellular material from swabs assessed using two samples from each cellular load and two independent production batches (Supplemental Table S3).

Functional verification of manufactured cellular input levels was also confirmed using the Genedrive® platform. Low-cell load materials were manufactured at approximately the assay LoD, while medium and high-cell load materials were manufactured at approximately 3× and 10× the LoD, respectively (Table 1). Dilution of the medium and high-cell load materials by factors of 3 and 10 yielded equivalent performance to the low-cell load materials, confirming that the intended relative cellular inputs had been achieved during manufacture. Across all materials and genotypes tested (n = 20), Genedrive® calls were fully concordant. These studies confirmed that materials generated at different target cellular loads exhibited the expected functional performance characteristics. Batch release subsequently supported by AQL-based attribute testing of the lowest cell-load material, using predefined defect criteria and random sampling plans (Table 2). All tested batches met the predefined acceptance criteria, with no defective swabs identified in the sampled units.

**Table 2.** Acceptable Quality Limit Testing (Level II) on *MT-RNR1* cell-based swab materials.

| Batch number* | Genotype | Batch Size | Swabs Tested | Swabs meeting Acceptance Criteria |
| --- | --- | --- | --- | --- |
| 1 | *MT-RNR1* m.1555G | 250 | 32 | 32 |
| 2 | *MT-RNR1* m.1555A | 135 | 17 | 17 |

\*Numbers are used for identification within this study; full manufacturing batch records are maintained by EMQN.

### MT-RNR1 Material Performance Under Procedural Variation

To assess the response of the manufactured swab materials to procedural variation, controlled modifications were introduced at key stages of the Genedrive® testing workflow (27) using low-cellular load swab materials. Altered cell-release procedures a limited effect on assay signal characteristics. Omission of swab rotation during cell release resulted in a significant reduction in peak height for the *m.1555G* genotype, whereas no significant effect was observed for the *m.1555A* genotype (Figure 2A). Despite reduced signal intensity, all samples produced genotype-specific melting curves with Tm values consistent with the expected *MT-RNR1* genotype (Supplemental Fig. S2A). In contrast, variation in lysate transfer volume had minimal impact on either peak height or Tm values (Figure 2B, Supplementary Fig. S2B).

**Figure 2.**
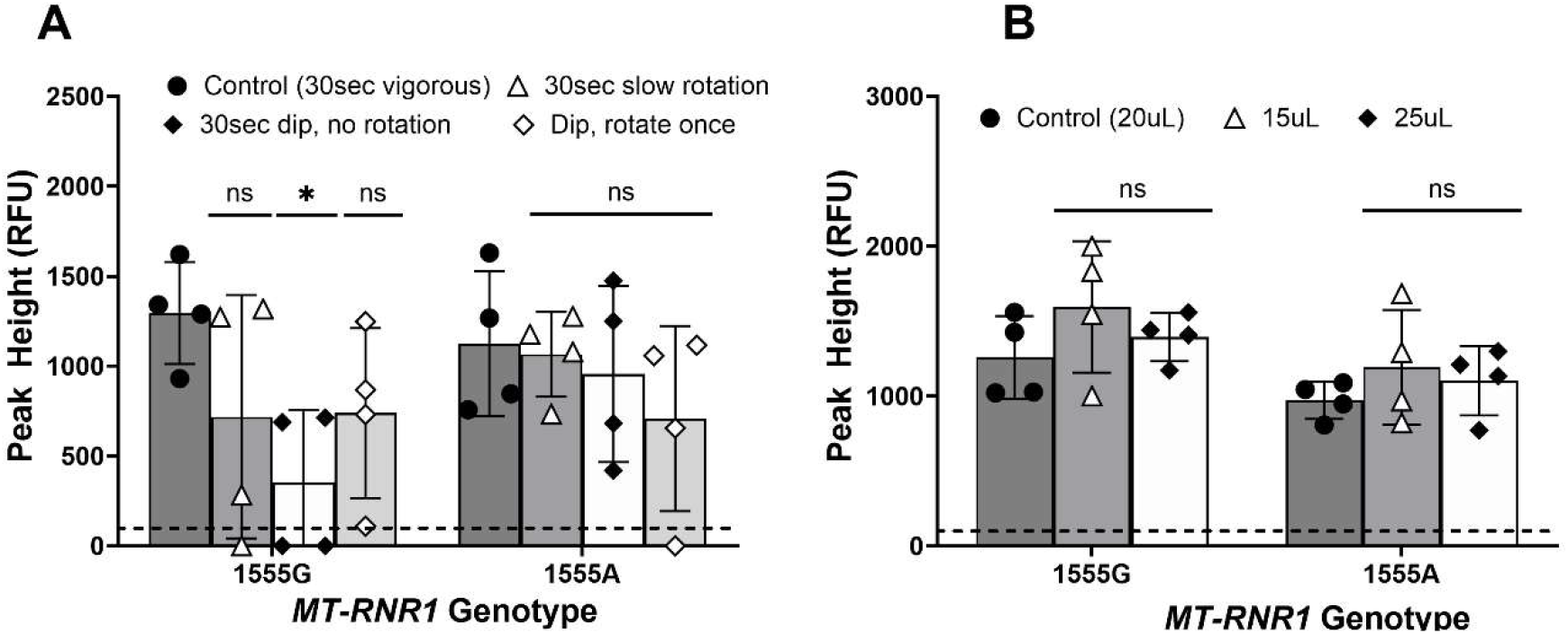
Cell release robustness of *MT-RNR1* swab-based reference materials. **(A)** Effect of alternative cell-release procedures on assay performance. The manufacturer’s recommended swab release procedure was compared with reduced-rotation and no-rotation conditions to simulate operator-dependent variation. **(B)** Effect of lysate transfer volume on assay performance. The manufacturer’s recommended lysate transfer volume was compared with reduced and increased transfer volumes to simulate deviations from the recommended testing procedure. Data are presented as mean ± standard deviation from independently processed swab samples. Peak heights are reported in Relative Fluorescence Unit (RFU). The dashed line indicates the peak height cut-off of 100 RFU. Statistical comparisons were performed using two-way ANOVA with Dunnett’s multiple-comparison test (ns, P > 0.05; *, P 0.01 to 0.05).

### Stability for storage, shipment, and in-use handling

Real-time stability studies demonstrated that the lower cell-load *MT-RNR1* swab materials remained stable for at least 2.5 years from the date of manufacture when stored at 2–30°C. Accelerated stability studies further supported an estimated shelf life of up to four years at 30°C. Shipment stability studies demonstrated maintenance of the expected genotype assignments following road and air transportation conditions representative of routine EQA distribution. In-use stability studies showed concordant genotype assignments and comparable assay signal characteristics following pouch opening and exposure to both ambient and elevated temperature and humidity conditions (Supplemental Table S4; Supplemental Figure S4)

### Clinical Verification During PALOH-UK Implementation

Five sites participating in the PALOH-UK study used the cell-based swab materials to clinically verify Genedrive® systems prior to service implementation. Across 11 Genedrive® systems, a total of 234 verification runs were performed. Of these, 232 (99.1%) generated valid results concordant with the expected genotype classifications across both *MT-RNR1* positive and wild-type specimens. Two test failures were reported, with no incorrect genotype assignments observed (Supplemental Table S5).

### Outcomes of the First MT-RNR1 PGx POCT EQA one year cycle

In the first year of programme operation, participation evolved across seven consecutive programme rounds shown in Figure 3A. Initial registration was driven primarily by sites involved in the PALOH-UK implementation study, reflecting the early UK rollout of *MT-RNR1* POC testing. Later rounds included additional participants from outside the PALOH-UK programme, including laboratories from Scotland, the Netherlands and the United States.

**Figure 3.**
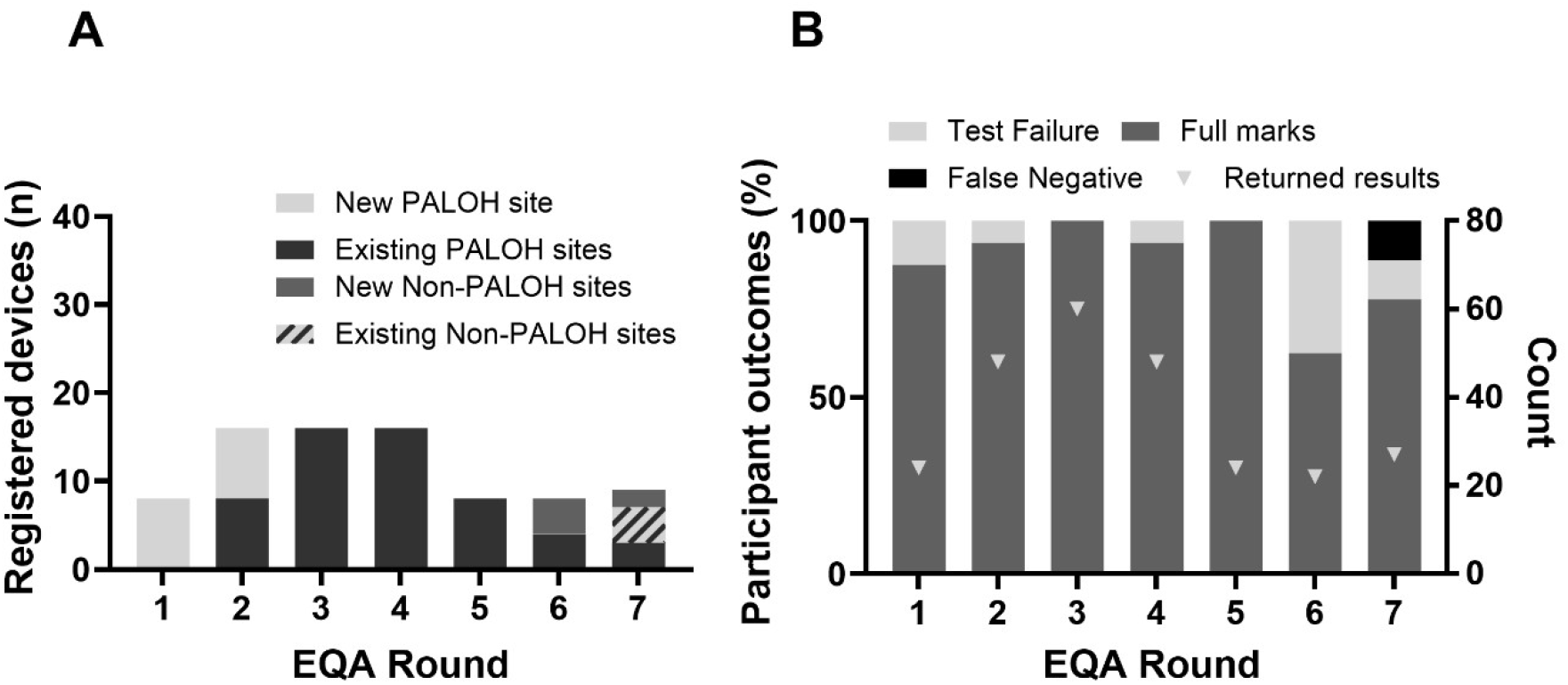
Uptake and performance of the *MT-RNR1* pharmacogenomic POCT EQA programme during the first year of implementation. **(A)** Number of registered devices across seven consecutive EQA rounds, stratified according to PALOH-UK associated and non-PALOH-UK sites and by new or returning participation status. Initial registration was driven primarily by sites participating in the PALOH-UK study, while later rounds demonstrated continued uptake from both PALOH-UK and non-PALOH-UK sites. Diamonds indicate the number of results returned for each EQA round (right y-axis). Survey 3 included an optional EQA sample. 5% to 100%. Test failures were infrequent and were associated with samples manufactured at cellular loads close to the assay limit of detection. One participating device reported a false negative.

Participation increased from eight registered devices in the first survey to a maximum of sixteen devices in subsequent rounds. Across the seven surveys, a total of 253 results were returned, providing one of the first systematic assessments of real-world performance for *MT-RNR1* pharmacogenetic POCT. The number of expected and returned results matched across all survey rounds, indicating that all distributed EQA samples were successfully tested and reported (Figure 3A).

Overall participant performance remained high throughout the assessment period (Figure 3B). Full-mark rates ranged from 87.5% to 100%, with only one device incorrectly reporting an *MT-RNR1* m.1555G-positive specimen as not detected (false negative). A total of six test failures were reported across the one-year cycle. Notably, five failures occurred with EQA specimens manufactured at low cellular input levels close to the assay’s established LoD, with the remaining failure occurring in a medium cellular input specimen. These more challenging materials were intentionally included to evaluate workflow robustness and are inherently more susceptible to variation arising from sample preparation and operational testing procedures.

## Discussion

Despite increasing clinical interest in PGx POCT, widespread implementation remains limited by the absence of quality-assurance approaches capable of supporting decentralised genomic testing outside specialist laboratories. Prior to this work, dedicated EQA programmes specifically designed for pharmacogenomic POCT were not available, and suitable workflow-commutable reference materials were lacking. The present study addresses this gap through the development of cell-based swab materials specifically designed to assess the complete PGx POCT workflow, from sample processing through to genotype assignment. Importantly, the framework was evaluated during active clinical implementation through the PALOH-UK programme, demonstrating how quality assurance can be embedded within service deployment rather than introduced retrospectively.

The key finding of this study is that fixed lymphoblastoid cell lines can be used to generate scalable, reproducible and homogeneous reference materials that closely mimic clinical specimens while supporting large-scale quality-assurance activities. Unlike extracted DNA controls, the developed materials challenge the complete testing pathway, including cell release, amplification and genotype assignment. The inclusion of materials spanning the operational range of the assays, including specimens positioned close to the LoD, enabled assessment of workflow robustness under realistic testing conditions. As expected, materials manufactured below the LoD exhibited an increased frequency of test failures associated with insufficient cellular input. However, when amplification signals met assay acceptance criteria, genotype assignments remained correct and clustered within the expected genotype-specific melting temperature ranges. This suggests that the reduced performance observed at very low cellular inputs was attributable to specimen quantity rather than impaired genotype discrimination. Importantly, these findings support the suitability of the selected manufacturing specifications and demonstrate that the materials accurately reflect assay performance at and around the analytical detection threshold. In addition, the materials were robust to procedural variation. Changes affecting cell release influenced signal intensity to a greater extent than variation in lysate transfer volume, although expected genotype assignments were maintained throughout. These findings suggest that efficient cellular recovery is likely to have a greater impact on test performance than modest deviations in lysate transfer volume.

Independent qPCR and functional testing confirmed that the manufactured materials reliably reproduced the intended differences in cellular input. This reproducibility is critical for EQA applications, ensuring that observed performance differences reflect testing performance rather than variation in sample manufacture.

The materials also demonstrated characteristics required for routine deployment. Real-time and accelerated stability studies supported long-term storage, while shipment and in-use studies confirmed suitability for international distribution and routine operational use. Together, these findings demonstrate the operational robustness required for EQA applications, where materials must maintain performance during storage, transport and routine laboratory handling.

The translational value of the approach was demonstrated through real-world implementation of the developed materials. In addition to supporting EQA, the materials were successfully used for clinical verification of 11 Genedrive® systems across five PALOH-UK participating sites prior to service implementation. Across 234 verification runs, 232 (99%) generated valid results concordant with the expected genotype classification demonstrate the suitability of the materials for supporting local verification of pharmacogenomic POCT services before clinical deployment.

Following implementation, the materials were deployed within the *MT-RNR1* PGx POCT EQA programme. Participation increased during the first year of operation, extending beyond the original PALOH-UK cohort to include laboratories from multiple countries. This expansion beyond the initial implementation sites suggests growing interest in pharmacogenomic POCT quality assurance and demonstrates the practical feasibility of delivering EQA for genomic point-of-care testing across diverse healthcare settings. Participant performance remained consistently high, with only six test failures and a single false-negative result reported across the first year of operation. Notably, most failures occurred in specimens manufactured at cellular input levels close to the assay LoD, supporting the inclusion of borderline-input materials within EQA schemes to assess workflow performance under more challenging testing conditions. These findings demonstrate that quality-assured PGx POCT can be implemented successfully outside traditional molecular laboratories when supported by appropriate reference materials and external assessment.

This study has several limitations. The first year of the EQA programme primarily involved UK-based sites and focused on a single POCT platform. Additional studies involving larger participant cohorts, diverse healthcare settings and multiple testing technologies will be required to establish broader applicability. Nevertheless, the data presented here demonstrate a scalable approach for supporting implementation, clinical verification and ongoing quality assurance of genomic POCT.

The principal contribution of this work is not solely the development of contrived materials, but the demonstration of an integrated framework that combines workflow-commutable reference materials, clinical verification and external quality assessment to support implementation of pharmacogenomic POCT. The successful implementation of the first *MT-RNR1* PGx POCT EQA programme provides a practical model for quality-assured deployment of genomic POCT and offers a generalisable framework for future pharmacogenomic and genomic POCT applications.

## Data Availability

All data produced in the present study are available upon reasonable request to the authors

## Acknowledgements

The authors would like to thank Sian Hilton for providing regular updates on the PALOH-UK study and for facilitating engagement between EMQN and NHS Trusts participating in the study. The authors also thank Victoria S Williams for her insightful discussions, constructive feedback, and continued support throughout the project.

## Research Funding

This work was supported by Innovate UK (10058536). JHM and WN are supported by the National Institute for Health and Care Research (NIHR) under its Invention for Innovation (i4i) Programme (NIHR208090), the NIHR BRC:Manchester (NIHR 203956) and the NHS England Genomics Programme through the Pharmacogenomics and Medicines Optimisation Network of Excellence. The views expressed are those of the author(s) and not necessarily those of the NIHR or the Department of Health and Social Care.

## Author Contribution

**S.P. and G.M.** conceived the concept of the artificial cell-based reference materials. **S.P., G.M. and S.A.** designed the cellular loading strategy. **G.M. and S.A** designed and performed the robustness study. **R.M.** conceived and led the study, coordinated the experimental programme, performed data analysis, interpreted the findings, and wrote the original manuscript draft. **S.P., R.M. and H.T**. designed the external quality assessment programme. **H.T.** contributed to study design, performed experimental studies, and assisted with data analysis and contributed to manuscript preparation. **JHM. and W.N**. provided patient samples for cell line generation, performed validation studies, and led the PALOH-UK implementation study.

All authors contributed to data interpretation, critically reviewed the manuscript, and approved the final version for publication.

## Supplemental Method S1

### Sanger sequencing

#### Polymerase chain reaction (PCR)

Primers were designed covering the genomic region for gene *MT-RNR1* using Primer3 design software available at http://primer3.wi.mit.edu/ and SNP checked. Primers pairs used in this study include:

##### For PCR, N13 tagged primers

- MT-RNR1 fwd 5’-gtagcgcgacggccagtCACGGGAAACAGCAGTGAT-3’
- MT-RNR1 rev 5’-cagggcgcagcgatgacGCGCCAGGTTTCAATTTCTA-3’

##### For Sequencing used N13 Primers

- N13 fwd 5’-GTAGCGCGACGGCCAGT-3’
- N13 rev 5’-CAGGGCGCAGCGATGAC-3’

Saliva samples containing ∼ 10ng – 15 ng genomic DNA were amplified using touchdown PCR programme in a 10 µl reaction consisting of 1x GoTaq® Hot Start Green Master Mix (Promega), 0.5 µM forward and reverse N13 linked *mtrnr1* primers (*Sigma Aldrich*). Reactions were cycled on a Veritii Pro Thermal Cycler (*Thermo-Scientific*).

#### Automated DNA sequencing

PCR products were purified using Agencourt Ampure XP Beads (Beckman Coulter). Essentially to 10 µl of PCR product 20 µl of Agencourt Ampure XP Beads was added and put through the Beckman Coulter multichannel robotics for the automation of the Agencourt AMPure protocol. This technique uses Solid Phase Reversible Immobilization (SPRI®) magnetic bead technology and a specifically optimized buffer to selectively bind PCR amplicons to paramagnetic beads whilst reaction contaminants such as excess primers, primer dimers, dNTPs, salts and enzymes are removed by a simple washing procedure. The resulting purified DNA was then sequenced. For sequencing reactions 1.8 µl of purified PCR product was added to a 10 µl sequencing reaction containing 0.25µl BigDye Terminator Mix v3.1 Cycle Sequencing Kit (Applied Biosystems), 1.875 µl of 5x Sequencing Buffer (Applied Biosystems), 0.5 μM forward or reverse N13 primer. Reactions were cycled 25 times using the following conditions: Initial denaturation at 96°C 1 minutes followed by 96 °C 10 sec, 52°C 5 secs, 60 °C 75 secs. DNA was then purified using the Beckman Coulter NX multichannel robotics for the automation of the Agencourt CleanSEQ 384 protocol. To 10 µl sequencing reactions 5 µl of CleanSEQ Beads were added (Beckman Coulter) and then put through the automated protocol using the Agencourt Solid Phase Reversible Immobilization Chemistry to remove unincorporated dye-terminators, nucleotides, salts and other contaminants prior to delilneation on capillary electrophoresis instrumentation. Purified sequencing reactions were then run on an ABI 3700 Sequencer.

#### Sequencing analysis

Patient sequences were compared to a wildtype control sequence using the Staden package Pregap4 1.4b1 and genotypes recorded.

## Supplemental Method S2

### Quantitative PCR (qPCR)

#### Design of Primers

Cells from artificial swabs containing high and low cell loads for were released according to the Genedrive® MT-RNR1 ID Kit IFU. To enable quantification irrespective of genotype, the housekeeping gene 18S rRNA (NC_000021.9) was selected as the target due to its stable expression and widespread use in qPCR applications (28). Primers were synthesised by ThermoFisher Scientific (Custom DNA Oligo Service; Cat. No. 10336022) and were based on sequences previously described by Shoop et al (29). The primer sequences were:

- Forward: 5’ CGGACAGGATTGACAGATT 3’
- Reverse: 5’ CTATTGCTTGCTCTGAGACC 3’

#### Sample Preparation

Cells from artificial swabs containing high and low cell loads for MT-RNR1. The resulting cell suspensions were used directly, without further dilution, as template material for the qPCR reactions described in Section 2.7.4.

#### Standard curve

Human Genomic DNA (Sigma-Aldrich, Catalogue number – 69237-3) was diluted in TE Buffer (Invitrogen, Catalogue number – 12090015) to a concentration of 10ng/µL. A five-point, 10-fold serial dilution series was then prepared, covering a DNA input range of 50 ng to 0.005 ng per reaction. Standard curve reactions were set up in the same method described in Section S2.4

#### Sample Quantification

Each reaction was prepared in a final volume of 20 µL and contained 1× PowerUp™ SYBR™ Green Master Mix (Applied Biosystems, Catalogue number – A25918), 0.5 µM each of the forward and reverse primers (ThermoFisher Scientific, Catalogue Number - 10336022), 5 µL of template DNA, and nuclease-free water (Sigma-Aldrich, Catalogue Number - W4502) to a final volume of 20 µL.

qPCR amplification was performed using a Bio Molecular Systems Mic Real-Time qPCR Cycler (4-channel). The cycling protocol comprised an initial denaturation step at 95°C for 5 min, followed by 40 cycles of denaturation at 95°C for 5 s, annealing at 60°C for 15 s, and extension at 72°C for 10 s. Samples, standards, and no-template controls were analysed in technical triplicates and at least two biological replicates. DNA concentrations in swab-derived samples were calculated by interpolation from a standard curve generated using human genomic DNA standards (section 2.7.3).

#### Statistical Analysis

All relevant data were exported using Workbench software v2.4.1 (Bio Molecular Systems, Queensland, Australia) and subsequently analysed using Microsoft Excel. The mean and standard deviation were calculated for each sample. Unknown samples were quantified based on their Cq values using the generated standard curve. The amplification efficiency, calculated from the standard curve slope, was 97.26%, and the linearity of the assay was demonstrated by an R² value of 0.9935.

Artificial swab materials were manufactured across four defined cellular input levels (define) to represent both routine testing conditions and challenge specimens positioned near the analytical limits of the corresponding Genedrive® MT-RNR1 ID Kit. Fixed-cell suspensions derived from the characterised LCLs were deposited onto flocked swab matrices to mimic clinical specimens and generate genotype-specific materials encompassing clinically relevant MT-RNR1 variants.

Fixed cell suspensions were used to manufacture flocked swab-based materials designed to simulate clinical sample collection workflows. Target cell concentrations were informed by both the analytical sensitivity of the Genedrive® *MT-RNR1* Kit and cell counts measured in clinically derived buccal swab specimens. Assay-specific limits of detection (LoD) were converted to equivalent cell numbers per swab using the Genedrive® sample preparation workflow; however, as these values assume complete cell recovery and transfer, manufacturing targets were selected to span clinically relevant cellular inputs observed in patient samples while remaining above the minimum analytical detection threshold. Swab manufacture was performed by Microbix Biosystems Inc. (Toronto, Canada) using validated proprietary processes to deposit fixed cells onto flocked swab matrix at predefined target cell loads. Manufacturing batches comprised a single production lot of at least 135 swabs of the same genotype and cellular input level. Swabs were manufactured to a target cell load within ±10% of the specified value. Manufactured swabs were individually packed in foil pouches and were stored at +4–18°C prior to distribution.

## Supplemental Method S3

### Stability Studies

Accelerated stability testing was conducted to predict long-term storage performance and support an estimated shelf life of up to four years at room temperature. Shipment stability was assessed using low cellular load manufactured swab materials distributed internationally by air and road transport using standard EMQN International Air Transport Association (IATA) compliant packaging (24). Temperature and humidity were monitored throughout transit. Samples from shipments representing the highest (33.6 °C, Canada) and lowest (26 °C, Germany) recorded temperatures were selected for analysis. Following return to EMQN, materials were stored at 4 °C until testing with the POCT Genedrive® ID Kit. Stability was assessed by comparison of observed and expected genotype assignments.

Out-of-pouch stability was also assessed to determine material performance following package opening. At predefined intervals, materials were analysed using the Genedrive® *MT-RNR1* and *CYP2C19* ID kits, and performance was evaluated based on genotype assignment.

## Supplemental Tables and Figures

**Table S1.** Clinical verification protocol used during the PALOH-UK study for Genedrive® MT-RNR1 systems, developed in accordance with ISO 15189:2022 and CLSI EP12. Primary verification was performed for the first Genedrive® system introduced into clinical service at a site, while secondary verification was performed for any additional Genedrive® systems.

| Primary Verification |  |  |  |  |
| --- | --- | --- | --- | --- |
| Sample | No. samples per day | Operator 1 days | Operator 2 days | Total tested samples |
| *MT-RNR1* WT (High cell load) | 2 | 1 & 3 | 2 & 4 | 8 |
| *MT-RNR1* WT (Low cell load) | 2 | 1 & 3 | 2 & 4 | 8 |
| *MT-RNR1* 1555G (High cell load) | 2 | 1 & 3 | 2 & 4 | 8 |
| *MT-RNR1* 1555G (Low cell load) | 2 | 1 & 3 | 2 & 4 | 8 |
| **Total** |  |  |  | **32** |
| Secondary Verification |  |  |  |  |
| Sample | No. samples per day | Operator 1 days |  | Total tested samples |
| *MT-RNR1* WT (High cell load) | 2 | 1 & 3 |  | 4 |
| *MT-RNR1* WT (Low cell load) | 2 | 1 & 3 |  | 4 |
| *MT-RNR1* 1555G (High cell load) | 2 | 1 & 3 |  | 4 |
| *MT-RNR1* 1555G (Low cell load) | 2 | 1 & 3 |  | 4 |
| **Total** |  |  |  | **16** |

**Note:** Primary verification was performed by two operators across four days. Secondary verification was performed by a single operator across two non-consecutive days on an additional Genedrive® system. Numbers represent individual test runs.

**Table S2.** Performance of cell-based swab material at the lowest intended manufacturing concentration.

| Gene | Genotype | Swabs Tested (n) | Expected genotype call (n) | Concordance |
| --- | --- | --- | --- | --- |
| *MT-RNR1* | m.1555A | 7 | 7 | 100% |
|  | m.1555G | 7 | 7 |  |
| **Total** | **-** | **14** | **14** | **100%** |

**Table S3.** Verification of cellular input levels in *MT-RNR1* cell-based swab materials.

| **Assay** | **Swab cellular load** | **DNA recovery (ng/swab) mean ± SD\*** | **Relative to low fold** | **Concordance with expected assay call (%)** |
| --- | --- | --- | --- | --- |
| *MT-RNR1* | High | 16.3 ± 4.7 | 9.1±2.9 | 100 |
|  | 10x high* | — | — | 100 |
| | Low | 1.8 ± 0.3 | 1 | 100 |

\*Cell released in the assay buffer diluted 10x
\*Calculated from two samples per cellular load across two independent *MT-RNR1* batches.

**Table S4.** Stability assessment of *MT-RNR1* cell-based swab materials.

| **Study** | **Condition** | **Duration (days)** | **Endpoint** | **Samples tested (n)** | **Outcome** |
| --- | --- | --- | --- | --- | --- |
| Accelerated stability | +50 °C (Arrhenius model) | 203 | Genotype call | 3 | 100% concordance |
| Shipment stability | International Air transport (+33.6 °)* | 9 | Genotype call | 3 | 100% concordance |
| Shipment stability | International Road transport (+26 °C)* | 6 | Genotype call | 3 | 100% concordance |

\*Maximum temperature recorded during shipment.

**Table S5.** Clinical verification of Genedrive® MT-RNR1 systems using cell-based swab materials during implementation of the PALOH-UK study.

| **Site** | **Genedrive System \*** | **Verification type** | **Verification run** | **Correct genotype call** | **Test failures** | **Success rate %** |
| --- | --- | --- | --- | --- | --- | --- |
| Site 1 | 1 | Primary | 32 | 32 | 0 | 100% |
|  | 2 | Secondary | 16 | 16 | 0 | 100% |
|  | 3 | Secondary | 16 | 16 | 0 | 100% |
| Site 2 | 1 | Primary | 32 | 32 | 0 | 100% |
|  | 2 | Secondary | 16 | 16 | 0 | 100% |
| Site 3 | 1 | Primary | 33 | 32 | 1 | 97% |
|  | 2 | Secondary | 16 | 16 | 0 | 100% |
| Site 4 | 1 | Primary | 32 | 32 | 0 | 100% |
|  | 2 | Secondary | 17 | 16 | 1 | 94% |
| Site 5** | 1 | - | 12 | 12 | 0 | 100% |
|  | 2 | - | 12 | 12 | 0 | 100% |
| **Total** |  | - | **234** | **232** | **2** | **99%** |

\*Numbers are used for identification within this study; corresponding device serial numbers are maintained by the PALOH-UK participating sites. \*\*Site 5 performed a reduced testing approach.

**Figure S1.**
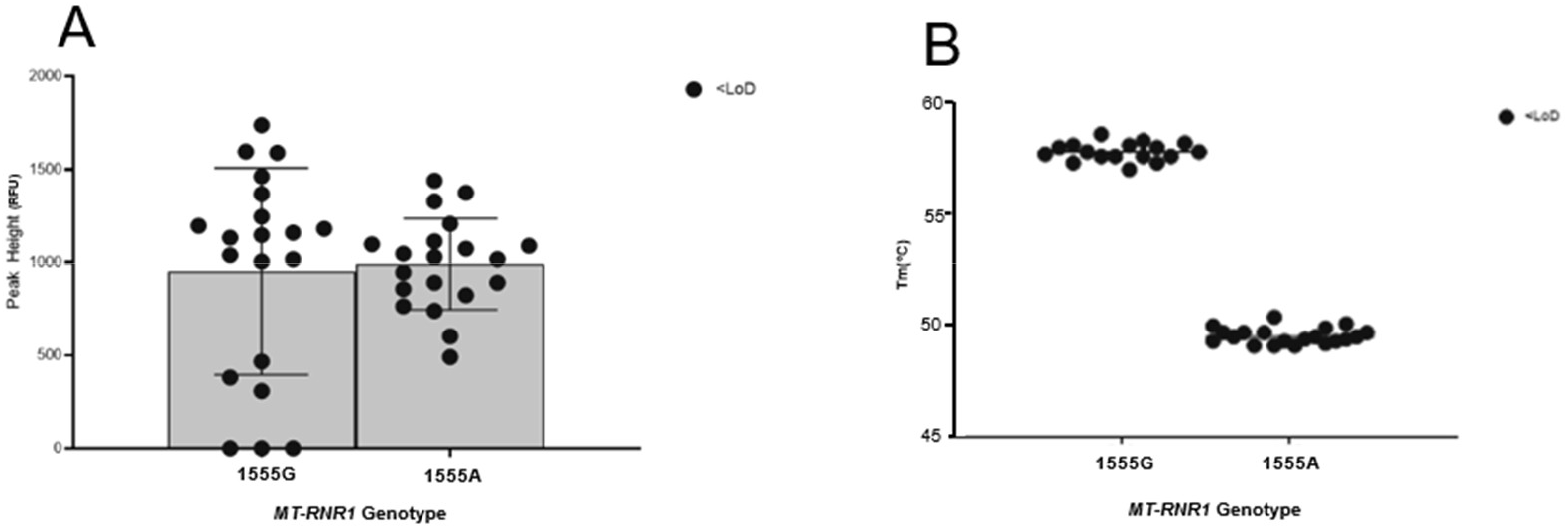
Assay peak-height and melting temperature distribution for cell-based swab materials manufactured below the assay LoD. Assay peak heights obtained from MT-RNR1 cell-based swab materials manufactured below the LoD. Corresponding melting temperatures (Tm) for samples generating a detectable peak signal are shown in the left panel. Samples producing sufficient amplification signal remained within the expected genotype-specific Tm cluster, supporting accurate genotype assignment despite reduced assay signal intensity. Data are presented as mean ± standard deviation from independently processed swab samples. Peak heights are reported in Relative Fluorescence Unit (RFU).

**Figure S2.**
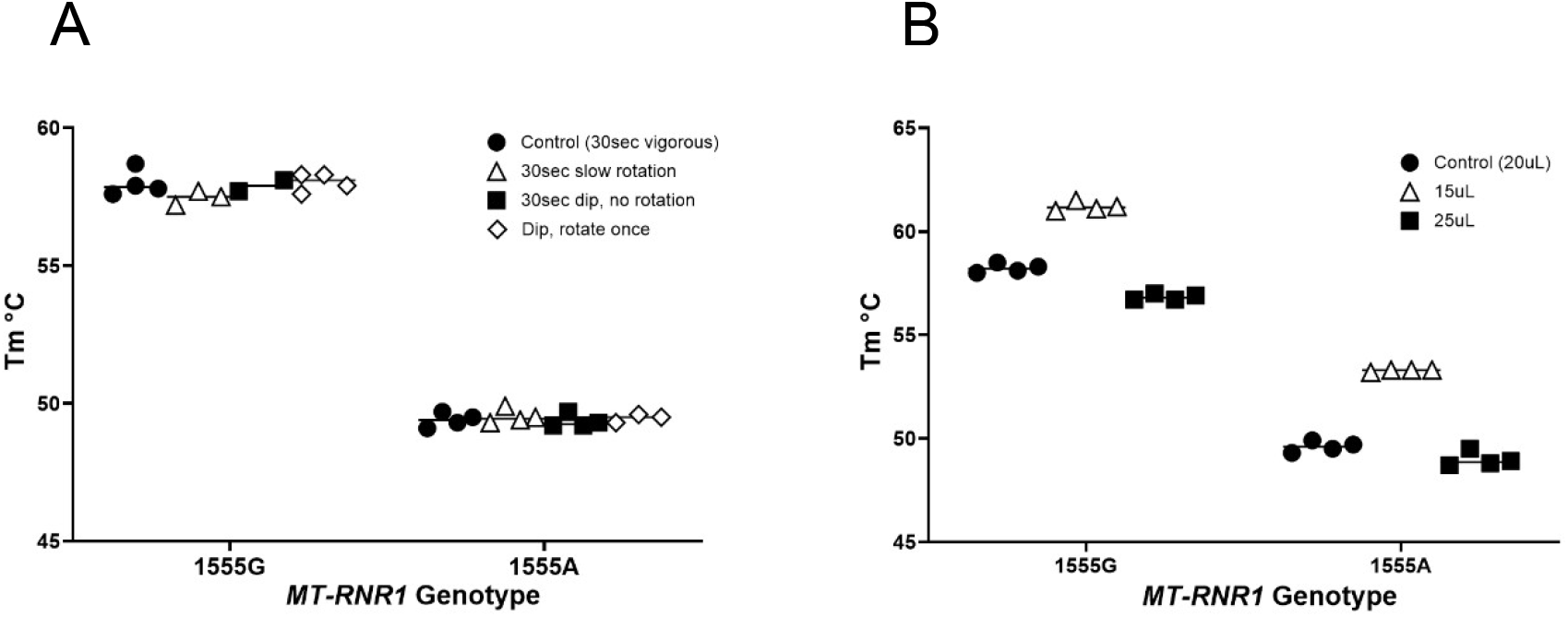
Stability of genotype-specific melting temperatures under workflow stress conditions. Melting temperatures (Tm) obtained using the Genedrive® *MT-RNR1* ID Kit cell-based materials following alternative cell-release procedures and variations in lysate transfer volume. Minimal variation in Tm was observed across conditions, and genotype-specific melting profiles were maintained throughout, confirming the robustness of genotype calling despite deviations from the standard workflow.

**Figure S3.**
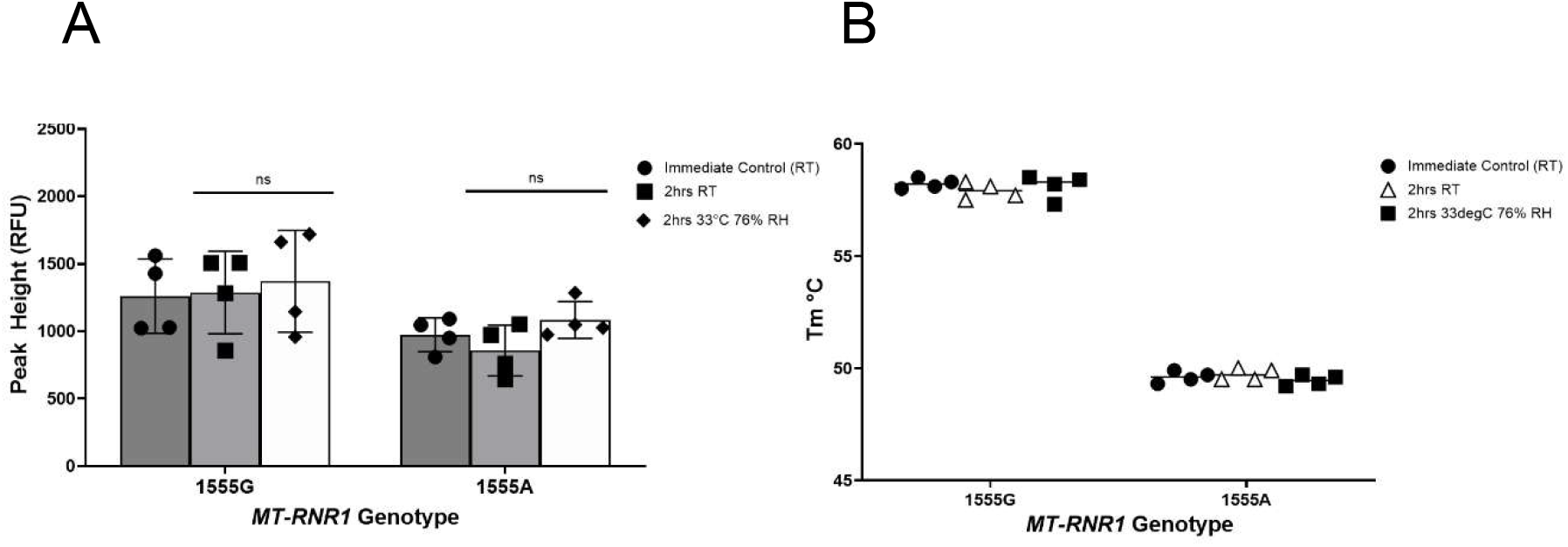
In-use (open-pouch) stability of *MT-RNR1* cell-based reference materials. Peak height values obtained using the Genedrive® *MT-RNR1* ID Kit for m.1555G and m.1555A swab materials following immediate testing, 2 h storage at room temperature (RT), and 2 h exposure to +37 °C and 70% relative humidity (RH) after opening. Comparable signal intensities and concordant genotype assignments were observed across all tested conditions. Data are presented as mean ± SD from *n* = 4 independently tested swabs. Statistical comparisons were performed using two-way ANOVA with Dunnett’s multiple-comparison test; ns, P > 0.05.

